# Perceived norms about male circumcision and personal circumcision status: a cross-sectional, population-based study in rural Uganda

**DOI:** 10.1101/2023.04.24.23288996

**Authors:** Jessica M. Perkins, Bernard Kakuhikire, Charles Baguma, Sehee Jeon, Sarah F. Walker, Rohit Dongre, Viola Kyokunda, Mercy Juliet, Emily N. Satinsky, Alison B. Comfort, Mark Siedner, Scholastic Ashaba, Alexander C. Tsai

## Abstract

**Introduction:** Over the past decade, 15 high-priority countries in eastern and southern Africa have promoted voluntary medical male circucmsion for HIV and STI prevention. Despite male circumcision prevalence in Uganda nearly doubling from 26% in 2011 to 43% in 2016, it remained below the target level by 2020. Little is known about perceived norms of male circumcision and their association with circumcision uptake among men.

**Methods:** We conducted a cross-sectional study targeting all adult residents across eight villages in Rwampara District, southwestern Uganda in 2020-2022. We compared what men and women reported as the adult male circumcision prevalence within their village (perceived norm: >50% (most), 10% to <50% (some), <10%, (few), or do not know) to the aggregated prevalence of circumcision as reported by men aged <50 years. We used a modified multivariable Poisson regression model to estimate the association between perceived norms about male circumcision uptake and personal circumcision status among men.

**Results:** Overall, 167 (38%) men < 50 years old were circumcised (and 27% of all men were circumcised). Among all 1566 participants (91% response rate), 189 (27%) men and 177 (20%) women underestimated the male circumcision prevalence, thinking that few men in their own village had been circumcised. Additionally, 10% of men and 25% of women reported not knowing the prevalence. Men who underestimated the prevalence were less likely to be circumcised (aRR = 0.51, 95% CI 0.37 to 0.83) compared to those who thought that some village men were circumcised, adjusting for perceived personal risk of HIV, whether any same-household women thought most men were circumcised, and other sociodemographic factors.

**Conclusions:** Across eight villages, a quarter of the population underestimated the local prevalence of male circumcision. Men who underestimated circumcision uptake were less likely to be circumcised. Future research should evaluate norms-based approaches to promoting male circumcision uptake. Strategies may include disseminating messages about the increasing prevalence of adult male circumcision uptake in Uganda and providing personalized normative feedback to men who underestimated local rates about how uptake is greater than they thought.

## INTRODUCTION

In 2007, the World Health Organization and the Joint United Nations Programme on HIV/AIDS recommended increasing voluntary medical male circumcision (VMMC) uptake in populations at high risk of HIV infection, especially in HIV-endemic countries [1]. This recommendation was based on research demonstrating male circumcision’s ability to reduce HIV acquisition risk among men [2-6] and indirectly decrease HIV acquisition risk among women [7, 8]. Male circumcision has also been associated with a reduced risk of acquiring other sexually transmitted infections such as syphilis, herpes simplex virus type 2, penile human papillomavirus, and cervical cancer [9-13]. Consequently, governments and international organizations began supporting efforts to enhance VMMC uptake [7, 14]. In 2016, the World Health Organization aimed to achieve an additional 25 million VMMCs among boys and men across 15 “high-priority” countries in eastern and southern Africa, including Uganda [15].

These efforts resulted in extensive VMMC uptake [6]. While less than 1.5 million VMMCs were recorded before 2012 in high priority countries, more than 26.8 million VMMCs were performed by 2019 [16]. However, uptake was less than needed to yield reductions in population-level HIV incidence [6]. Moreover, only about 50% of VMMCs in 2018 were conducted among men aged 15 years and up [16]. Thereafter, the World Health Organization emphasized increasing VMMC uptake rates within the general population in generalized HIV epidemic settings [17]. They also suggested targeting men at high risk of HIV acquisition and young men [17]. Subsequent data from 2020 indicated that about 75% of VMMCs in 2020 were among men aged 15 to 49 years with greater uptake among 15 to to 24 year olds compared to other age groups [16].

The Ministry of Health in Uganda implemented a Safe Male Circumcision Policy in 2010 [18, 19]. The male circumcision prevalence among men aged 15 to 49 years increased from 26% in 2011 to 43% in 2016-2017 [18, 19]. Moreover, the total number of circumcised men increased by 36-fold from 2010 to 2019, with more than 5.8 million boys and men circumcised by 2020 [6]. However, the male circumcision prevalence in Uganda remains lower than in other high priority countries [20, 21]. Increasing VMMC uptake requires novel implementation strategies, especially those that do not stigmatize uncircumcised individuals. Prior studies have found that some communications about VMMC suggest that uncircumcised men are less masculine than ‘real men’, ‘dirtier’ than circumcised men, and/or less virile/able to perform sexually [22]. This kind of messaging stigmatizes men choosing not to get circumcised or who do not have access to circumcision services. Stigmatizing messaging is ineffective at best and potentially harmful at worst [23]. However, focusing on actual local health-promoting norms about male circumcision represents an understudied potential strategy for increasing VMMC uptake.

### Conceptual Framework

Theoretical and empirical research has shown that perceived descriptive norms – that is, what individuals think most other people do – influence individuals’ own behaviors and beliefs [24-34]. Yet, individuals often misperceive local norms. They tend to underestimate the prevalence of peers who engage in health-promoting behaviors [35-45] and overestimate peers’ engagement in health-risk behaviors [35-45]. Health-promoting behaviors may be invisible due to associated stigma, local taboos preventing conversation, or because they are relatively private. Norm misperceptions arise due to lack of conversation or visible cues in the local environment about health-promoting behaviors, social media highlighting instances of high risk behavior, and biases in conversational, memory, and psychological inference processes [46, 47].

Altering perceived norms is an effective strategy for behavioral change [46-54]. Research across fields and topics has demonstrated that enhancing the salience and visibility of positive, health-promoting norms within local social networks or environments can shift individuals’ expectations about typical and acceptable behavior and attitudes. Consequently, individuals who had misperceived a risk behavior as the norm are less likely to engage in or initiate such behavior [55-66]. Furthermore, they may be less permissive of risk behavior in others, less likely to spread norm misperceptions in conversation, and more inclined to support others’ engagement in the desired behavior. This approach is particularly effective when individuals perceive information about local norms as recent, credible, and unaccompanied by contradictory or fear-based messaging [47, 49, 50]. Changing misperceived norms across a population can also influence collective attitudes and action [67]. Additionally, recent research has suggested that highlighting dynamic or trending norms, i.e., behaviors that are increasing in prevalence over time, can also promote behavioral change [68-71].

No studies have assessed perceived descriptive norms about male circumcision uptake in Uganda. Young uncircumcised men in eSwatini were more likely to report a personal intention to get circumcised if they thought that their friends, parents, or partner encouraged male circumcision and if they thought that most male friends were circumcised [72]. Similarly, young uncircumcised men in Zimbabwe were more likely to report a personal intention to get circumcised if they believed that their mother would encourage male circumcision and if they believed that male friends would get circumcised [73]. A recent systematic review concluded that familial and peer support for male circumcision facilitates uptake of VMMC [74]. Likewise, hearing about circumcision promotion from influential people such as religious leaders or a peer who underwent VMMC is also associated with circumcision uptake [75, 76].

Recent studies on HIV prevention, substance use, violence, and other health-related behaviors in Uganda and South Africa have found that individuals often overestimate the prevalence of community peers engaging in behaviors that increase the risk of HIV acquisition and transmission (e.g., avoiding testing, heavy substance use, condomless sex, intimate partner violence, and non-adherence to antiretroviral medication routines) [77-88]. Perceived norms about male circumcision in this context remain unexplored. Reducing HIV incidence in this setting necessitates greater VMMC adoption among men. Therefore, this study investigates men’s and women’s perceptions of the local male circumcision prevalence across eight villages in rural Uganda, comparing these perceptions to the actual circumcision prevalence among village men. Additionally, the study examines the relationship between perceptions and personal circumcision status among men aged 18 to 49 years. The findings will collectively indicate the potential for an opportunity to motivate VMMC uptake by addressing perceived norms, particularly in an HIV-endemic setting where numerous men in the general community are at high-risk for HIV acquisition.

## METHODS

### Study setting and design

We conducted a cross-sectional, whole-population study among all residents aged 18 years and older across eight villages in one rural, administrative parish in Rwampara District, southwestern Uganda. The study site was about 20 km from Mbarara City. The study team selected this parish in collaboration with local leaders, due to its tractable population and geographic size, and due to its similarity to other rural areas in Uganda where the majority of Ugandans reside [89]. Most households engage in an agriculture-based economy or small-scale trading/enterprise, household food and water insecurity are common, and access to electricity and piped water is rare [89-92]. These characteristics are similar to descriptions of other low-resource rural contexts in eastern and southern Africa. Moreover, this context is similar to rural areas in the 15 priority countries targeted for VMMC uptake in eastern and southern Africa.

### Study procedures

Research assistants who spoke the local language (Runyankore) gathered data in 2020-2022. All eligible individuals were invited to participate in this study if they were not incapacitated at the time of data collection. Using a continuously updated parish census list of eligible adult residents, research assistants contacted potential participants and asked them to participate in a study about health and wellbeing. During an informed consent process, research assistants obtained participants’ signature or thumbprint (for those unable to write). Research assistants recorded responses to a close-ended interview survey using a computer-assisted tool. The survey questions were written in English, translated into Runyankore, and then back-translated to English to verify the translation’s reliability. Question piloting and translation followed an iterative process. If procedures could not be conducted in person (typically around a participant’s home) due to coronavirus-19 restrictions, then the research assistants conducted consent and data collection over the phone. Study participants received a kilogram of sugar or bar of soap (per local norms) for their time.

### Measures

One question elicited personal circumcision status from male participants. Response options included yes or no, though participants could also report don’t know or refuse to answer. A second question elicited estimates of the male circumcision prevalence in one’s own village (i.e., the perceived norm about male circumcision uptake) from both male and female participants. Specifically, each participant was asked how many men in their own village were circumcised, using a 5-point Likert-type scale ranging from ‘all or almost all men (>90%)’, ‘more than half of men but fewer than 90%’, ‘fewer than half of men but more than 10%’, and ‘very few or no men (<10%)’, and ‘do not know’. Pre-testing suggested that participants easily understood ‘Other adult men in your village’ as a reference group. Therefore, that group was set as the social reference group for identifying local norms. Other studies conducted in this setting have used similar wording to capture perceptions about local norms [93, 94]. The perceived norm response categories are subsequently referred to as ‘most’, ‘some’, ‘few’, and ‘don’t know’.

### Additional covariates

Past research has found that male circumcision uptake varies by sociodemographic characteristics [20, 95-102], HIV testing history [76, 100, 101, 103], knowing one’s HIV status [102], being HIV negative [102], and having condomless sex [95]. Thus, this study assessed several additional factors including perceived personal HIV risk/status (HIV-positive, no/low risk, medium/high risk (plus very few individuals with an unknown status)), having had condomless sex with a non-spouse sex partner in the past year, having had an STI in the past year, and having been tested for HIV in the past year. Sociodemographic variables included age, marital status (married/cohabiting versus divorced/separated/single), religion (Protestant, Catholic, Muslim, and Other), education (completed primary versus did not), household wealth quintile, and number of people in the household. To measure household wealth, we created a household asset index by conducting a principal components analysis on 26 separate variables representing household assets and housing characteristics (no missing data). We retained the first principal component to define the wealth index and then split it into quintiles [104, 105]. Finally, we created a variable representing whether any female household members thought that most men in their village were circumcised. We included this variable as a potential confounder between men’s perceived norms and men’s circumcision status given past work.

### Statistical analysis

We first calculated descriptive statistics of the population and the prevalence of male circumcision uptake among all men and among men <50 years old. Then we examined perceived norms about male circumcision uptake overall, among men, among women, and within subgroups across the population. For the subsample of men less than 50 years of age, we estimated the association between personally being circumcised and one’s perception about the male circumcision norm in his community. We fitted modified Poisson regression models; with a binary dependent variable, the modified Poisson regression model has been shown to yield estimated incidence rate ratios that can be interpreted straightforwardly as relative risk ratios [106]. The model adjusted for HIV perceived risk and status, history of HIV testing, any STI in past year, condomless sex with nonspouse partner in past year, age, marital status, education, wealth, religion, number of household members, and whether any women in the household thought most village men had been circumcised. We also conducted a sensitivity analysis to assess the extent to which the findings might change with the inclusion of all men in the sample (i.e., including men 50 years of age and older and men who identified as Muslim). Analyses were conducted with Stata version 16 and accounted for clustering of observations by village [107].

## RESULTS

Among 1723 people who were eligible for study participation, 1,566 were interviewed (90.9% response rate). The mean age across the full population was 42 years (standard deviation [sd] = 16). Most participants [940 (60%)] had completed primary education or more, and most were married/cohabiting as if married [1024 (65%)]. Twenty-two participants (1%) identified as Muslim. Forty-five percent (n=698) were men. Among the 472 men who were < 50 years old, the average age was 34 years (sd = 8). In this subpopulation, 217 (49%) had been tested for HIV in the past year, 73 (16%) had condomless sex with a non-spouse partner in the past year, 32 (7%) reported having an HIV-positive status, and 55 (13%) reported medium or high perceived HIV risk.

### The local prevalence of male circumcision

Among all male participants, 660 out of 698 men reported their circumsion status. Among the 38 who did not, 21 men who had never had sex were accidentally not asked about their personal circumcision status due to a logic branching error during data collection and 17 other men simply did report their circumcision status. Among the 660 men with a reported circumcision status, 191 (27%) men reported that they were personally circumcised. This prevalence ranged from 23% to 37% across the eight villages. All men who identified as Muslim reported being personally circumcised.

Among 444 male participants under 50 years old who provided a response about their circumcision status, 167 (38%) reported that they were personally circumcised, which ranged from 27% to 51% across villages. The male circumcision prevalence was higher among younger age groups. For example, almost half of men aged 18-25 years old were circumcised (n=40[48%]). The prevalence was lower among men who had not completed primary school (**Table 1**).

**Table 1.** Distribution of male study participants < 50 years old and the prevalence of male circumcision based on personal reports from all male residents aged 18 to <50 years old across eight villages in Rwampara District, southwest Uganda.

|  | N and percent of men less than 50 years old who reported their circumcision status<sup>a</sup> |  | n and percent of these men who reported being circumcised |  |
| --- | --- | --- | --- | --- |
| Total | 444 | 100% | 167 | 38% |
| Age (years) |  |  |  |  |
| 18-25 years old | 84 | 19% | 40 | 48% |
| 26-35 years old | 160 | 36% | 64 | 40% |
| 36-45 years old | 152 | 34% | 49 | 32% |
| 46-55 years old | 48 | 11% | 14 | 29% |
| Marital status |  |  |  |  |
| Not married | 147 | 33% | 57 | 39% |
| Married / cohabiting as if married | 297 | 67% | 110 | 37% |
| Religion |  |  |  |  |
| Catholic | 93 | 21% | 40 | 43% |
| Muslim | 7 | 2% | 7 | 100% |
| Protestant | 321 | 72% | 110 | 34% |
| Other (not religious, seventh day adventist) | 23 | 5% | 10 | 43% |
| Education |  |  |  |  |
| Less than primary education | 112 | 25% | 27 | 24% |
| Completed primary education | 332 | 75% | 140 | 42% |
| Household asset wealth |  |  |  |  |
| 1st (poorest) | 83 | 19% | 28 | 34% |
| 2nd | 99 | 22% | 36 | 36% |
| 3rd | 90 | 20% | 34 | 38% |
| 4th | 87 | 20% | 31 | 36% |
| 5th (least poor) | 85 | 19% | 38 | 45% |
| Lived with at least one woman who thought that >50% of men in village had been circumcised |  |  |  |  |
| No | 359 | 81% | 132 | 37% |
| Yes | 85 | 19% | 35 | 41% |
| Was tested for HIV in past 12 months |  |  |  |  |
| No | 227 | 51% | 74 | 33% |
| Yes | 217 | 49% | 93 | 43% |
| Had an STI in past 12 months |  |  |  |  |
| No | 411 | 93% | 153 | 37% |
| Yes | 33 | 7% | 14 | 42% |
| Had condomless sex with a non-spouse partner in past 12 months |  |  |  |  |
| No | 371 | 84% | 136 | 37% |
| Yes | 73 | 16% | 31 | 42% |
| Perceived personal HIV risk |  |  |  |  |
| HIV-positive | 32 | 7% | 9 | 28% |
| No/low risk | 353 | 80% | 137 | 39% |
| Medium/high risk | 55 | 13% | 20 | 36% |

<sup>a</sup>Circumcision status for 28 participants was unknown: 9 refused to answer and 19 were accidentally not asked the question after indicating that they had never had sex. They are not included in this table.

### Perceptions about the local male circumcision prevalence among adult men and women

Across all participants, 275 (18%) thought that most men in their villages had been circumcised (including 10 participants who thought that almost all men had been circumcised); 631 (40%) thought that some men in their villages had been circumcised; 366 (23%) thought that few men in their villages had been circumcised; and, 287 (18%) reported that they did not know how many men in their villages had been circumcised.

The overall pattern of perceived norm about local male circumcision uptake was similar between men and women, though more women indicated a ‘do not know’ response (**Figure 1**). The perceived norm that few village men had been circumcised varied in prevalence from 10% to 35% across sex-specific sociodemographic and HIV risk categories (**Supplemental Tables 1 and 2**).

**Figure 1.**
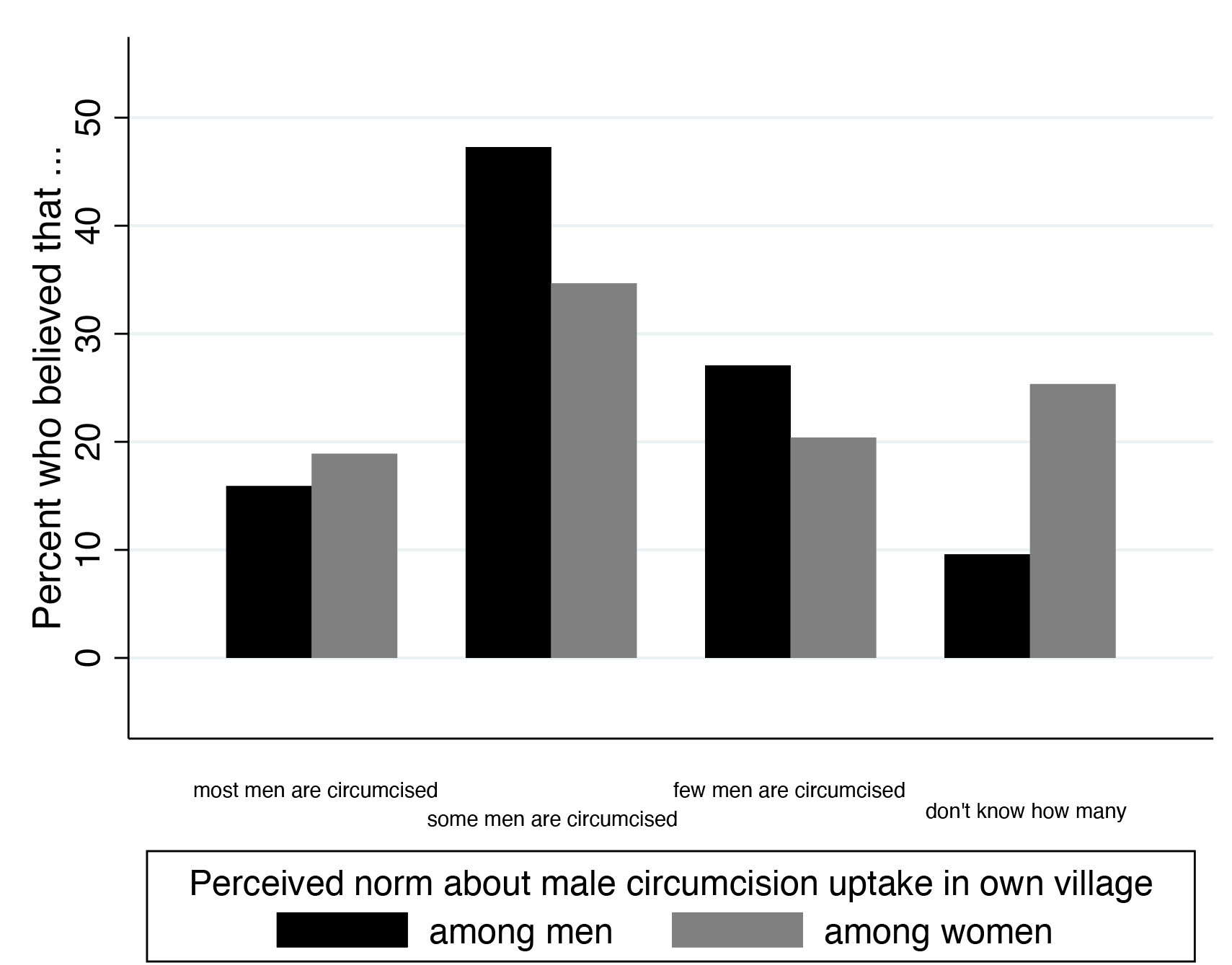
The distribution of perceptions about the prevalence of male circumcision in one’s own village among adults across eight villages in Rwampara District, southwestern Uganda (N=1566).

### Perceived norms as correlates of personal circumcision status among men <50 years of age

In the subset of men under 50 years of age and who did not identify as Muslim, we fitted a multivariable Poisson regression model specifying personal circumcision status as the dependent variable. Men who perceived that most men in their villages had been circumcised had a higher risk of having been circumcised than men who perceived that some men had been circumcised (adjusted relative risk [aRR] = 1.68; 95% CI, 1.22-2.30, p <0.001). Men who perceived that few men in their villages had been circumcised had a lower risk of having been circumcised (aRR = 0.51; 95% CI, 0.35-0.74, p <0.001). While the risk of having been circumcised appeared to decrease with age, estimates were not precisely estimated. Men who had finished primary education or more had a greater risk of having been cirucumcised, but the estimate was imprecise (aRR = 1.54; 95% CI, 0.98-2.42, p <0.064). None of the HIV risk factors nor any of the other sociodemographic factors were associated with participant circumcision status (**Table 2**). The predicted probabilities of being circumcised by perception categories are presented in **Figure 2**. Results from the sensitivity analysis including all men were similar (**Supplemental Table 3**).

**Table 2.** Modified multivariable Poisson regression model estimating associations between the perceived norm about circumcision status among men in one’s village and being personally circumcised among almost all resident adult men <50 years old (excluding Muslim men) across eight villages in Rwampara District, southwestern Uganda (n=433 men).

|  | Yes, circumcised |  |  |
| --- | --- | --- | --- |
|  | aRR | (95% CI) | p-value |
| Perceived norm about male circumcision uptake in own village |  |  |  |
| Most men are circumcised (i.e., >50%) | 1.68 | (1.25 – 2.27) | 0.001 |
| Some men are circumcised (i.e., 10% to <50%) | REF | - | - |
| Few men are circumcised (i.e., 0 to <10%) | 0.51 | (0.35 – 0.74) | <0.001 |
| Don't know how many men are circumcised | 0.59 | (0.33 – 1.06) | 0.079 |
| Age (years) |  |  |  |
| 18-25 | REF | - | - |
| 26-35 | 0.91 | (0.69 – 1.22) | 0.536 |
| 36-45 | 0.81 | (0.63 – 1.04) | 0.105 |
| 46-55 | 0.72 | (0.44 – 1.19) | 0.202 |
| Married / cohabiting (vs. other) | 1.17 | (0.91 – 1.50) | 0.216 |
| Religion |  |  |  |
| Catholic | REF |  |  |
| Protestant | 0.85 | (0.60 – 1.21) | 0.372 |
| Other | 1.10 | (0.63 – 1.90) | 0.743 |
| Completed primary education or more (vs. did not) | 1.54 | (0.98 – 2.42) | 0.064 |
| Household asset quintile |  |  |  |
| 1st quintile (poorest) | REF | - | - |
| 2nd quintile | 0.96 | (0.73 – 1.26) | 0.762 |
| 3rd quintile | 0.88 | (0.51 – 1.52) | 0.650 |
| 4th quintile | 0.86 | (0.57 – 1.28) | 0.449 |
| 5th quintile (least poor) | 1.18 | (0.77 – 1.80) | 0.451 |
| Lived with at least one woman who thought >50% of men in village had been circumcised (vs. did not) | 1.06 | (0.68 – 1.68) | 0.786 |
| Tested for HIV in past 12 months (vs. did not) | 1.18 | (0.80 – 1.74) | 0.413 |
| Had an STI in past 12 months (vs. did not) | 1.10 | (0.60 – 2.03) | 0.758 |
| Had condomless sex with a non-spouse partner in past 12 months (vs. did not) | 1.01 | (0.83 – 1.23) | 0.907 |
| Perceived personal HIV risk |  |  |  |
| HIV-positive | 0.88 | (0.58 – 1.35) | 0.572 |
| No/low risk | REF | - | - |
| Medium/high risk | 1.07 | (0.86 – 1.33) | 0.561 |

aRR = adjusted relative risk ratio, CI = confidence interval, REF = reference group

**Figure 2.**
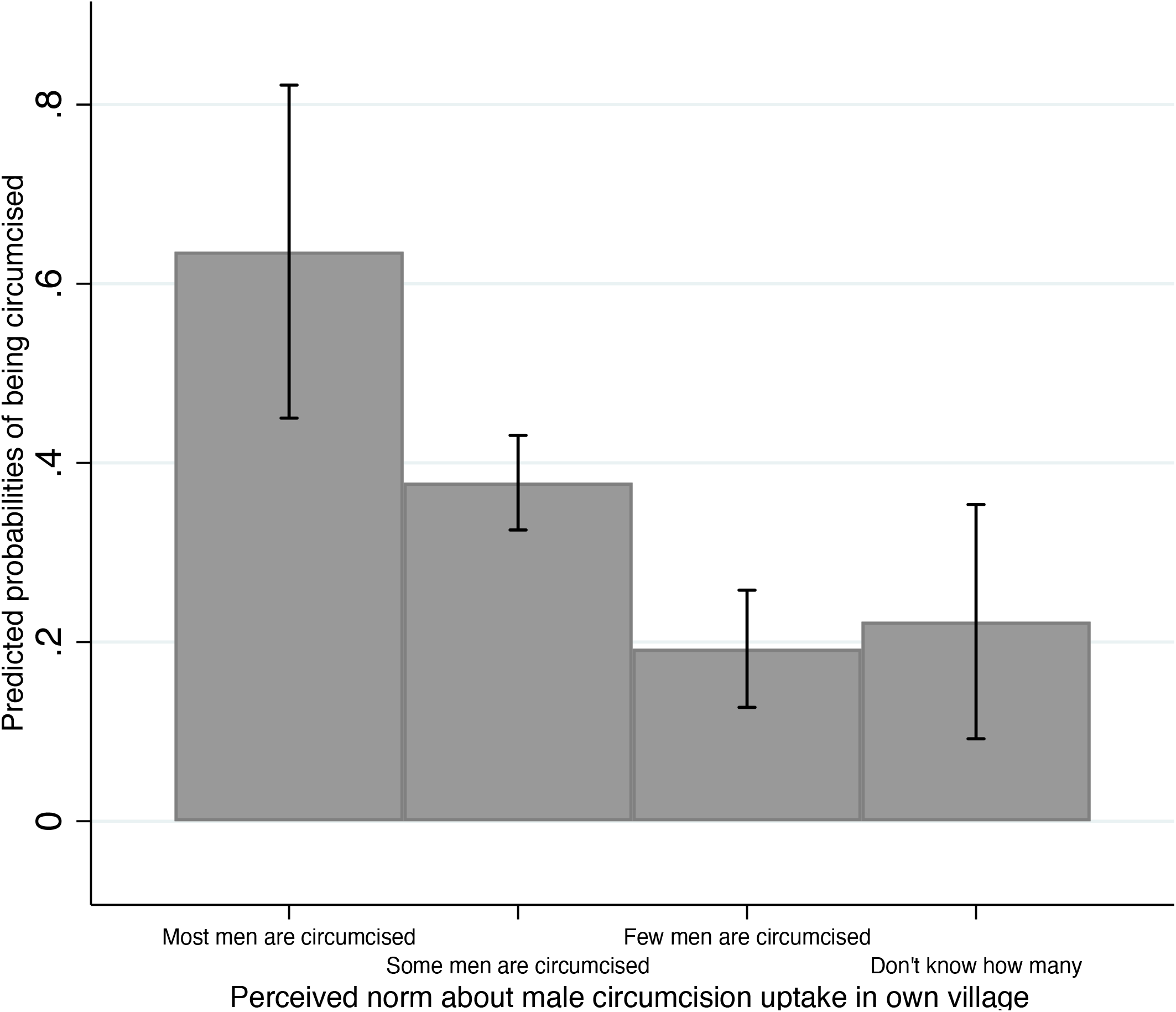
Predicted probabilities of being circumcised, stratified by their perception of the male circumcision prevalence in their own village after adjusting for other sociodemographic factors, among men <50 years of age across 8 villages in rural Uganda.

## DISCUSSION

This population-based study across eight villages in rural Uganda found that 38% of men aged 18 to 49 were circumcised (and 27% overall). However, 27% of men and 21% of women under-estimated the prevalence of circumcision among men in their villages. They incorrectly thought that few or no men were circumcised. Men who underestimated the prevalence of circumcision were less likely to be circumcised. These findings were robust and large in magnitude. Results about underestimating the prevalence of health-promoting behavior and the association between perceived norms and personal behavior are similar to findings from studies about other HIV-related issues such as HIV testing, adherence to antiretroviral therapy, condom use, and HIV-related stigma in this context and elsewhere in eastern and southern Africa [65, 77, 78, 81, 85].

Our results indicate the potential for employing a social norms approach to increase VMMC uptake in this context. This method, referred to as a ‘social norms approach’ [47], a ‘norms correction’ approach [108], or more broadly, a ‘norms-based’ approach, can be implemented by providing personalized normative feedback [109-112] about perceived versus actual circumcision rates to uncircumcised men and their partners who think few men are circumcised. A social norms campaign [56, 113, 114] emphasizing VMMC as a trending norm [71] in Uganda to the general population could serve as another strategy to correct underestimates and reinforce support for male circumcision uptake.

Addressing prevalence underestimates among uncircumcised men (i.e., correcting their state of false consensus [115]) may motivate them to undergo circumcision, either to conform with the growing trend or to seek ways to covercome barriers to getting circumcised. Increased awareness of the actual descriptive norm may also weaken negative beliefs about circumcision and encourage action on previous, unfulfilled intentions. Furthermore, rectifying prevalence underestimations among circumcised men (i.e., correcting their state of pluralistic ignorance [116]) may prompt them to share their experiences, realizing their behavior aligns with the trend. They might be more inclined to publically advocate for VMMC or support others in seeking options to do so. More information regarding the higher-than-expected and increasing circumcision prevalence could help mitigate stigmatizing beliefs or misconceptions, potentially arising from secrecy around the practice.

Correcting underestimates among women or a lack of information about male circumcision uptake and its upward trend may also promote male circumcision uptake. Mothers, sisters, and female partners often influence men’s circumcision decisions [72, 73, 117-120]. Changing their perceived norms may reduce personal stigmatizing attitudes toward male circumcision, increase conversation in support of circumcision, increase support for male partners to navigate any access barriers, and prevent spread of norm misperceptions. A qualitative study in Uganda found that pressure from female partners or wives to undergo circumcision also resulted in men undergoing VMMC [120]. Men reported that their partners believed in the importance of circumcision after hearing about it from radios, newspapers, and health services [120].

Examples of messages based on factual information rather than preferences, fantasies, or inaccurate beliefs include: “Recent data from your village indicates an increasing number of both single and married men choosing circumcision. In 2011, one in ten men were circumcised in Uganda, while by 2021, more than one in three single men and one in three married men in this village were circumcised”, and “The number men choosing to get circumcised continues to grow. Now almost half of men aged 18 to 35 in your village. If you are interested in learning more, please let me know. Male circumcision services are available here”. Health care providers and community health workers could share this information during routine visits or as part of other HIV- or contraception-related interventions (e.g., HIV testing and counseling visits, peer counseling, financial incentive programs, SMS-based adherence support, or couples-based support programs [121-126]). Local leaders could receive training on trending norms information, promoting male circumcision visibility and salience through facilitated discussions or one-on-one conversations [75]. Visual representations of local norms could be displayed at the entrances of clinics offering VMMC services [127], or publicised through social norms marketing such as radio or edu-entertainment messages [48, 64]. Future research should evaluate the effectiveness of such norms-based strategies on perceived norms and VMMC uptake. Continuous monitoring and assessment are crucial to ensure messages do not communicate stigmatizing attitudes or cause unintended consequences.

This study has limitations. First, because the data represent a single rural parish, findings may not generalize to the national population or to other countries. However, they do represent the entire parish population, and the study context is similar to rural areas across Uganda and in eastern and southern Africa. Thus, the findings provide a foundation for conducting research on perceptions about male circumcision norms in similar contexts where the prevalence of male circumcision has increased since initial WHO recommendations. Second, circumcision status is based on self-report. However, under-reporting and over-reporting of this behavior are likely to cancel each other out. Moreover, the slightly larger prevalence of circumcision among men 18 to < 50 years old in this parish in 2020-2022 (i.e., 30%) compared to the prevalence of circumcision among men aged 15 to < 50 years old in southwest Uganda in 2016 (i.e., 26%) [128] makes logical sense given the increasing male circumcision trend. Finally, the data are cross-sectional and we do not make claims of causality. Rather, we highlight opportunities where altering perceived norms about male circumcision by increasing the visibility of an actual upward trend might motivate subsequent behavior around VMMC uptake. Prior research has shown this type of norms-based strategy to be effective method of behavior change.

## CONCLUSION

In this population-based study in rural Uganda, approximately one in four adults underestimated the prevalence of male circumcision within their village. These perceptions were significant, as men’s underestimation of the circumcision prevalence was associated with a lower chance of being circumcised. A novel opportunity exists to promote VMMC uptake in HIV-endemic settings through a norms-based approach. Male circumcision in Uganda has trended upward since 2011. Providing information about the rising trend in male circumcision rates in this context and correcting prevalence underestimates will enhance VMMC uptake visibility and salience. This norms-based strategy to promote VMMC uptake could complement efforts to educate individuals about circumcision and its health benefits.

## Data Availability

All data produced in the present study are available upon reasonable request to the authors

## Authors’ Contributions

JMP conceptualized and designed the study. JMP and SJ wrote the first draft. BK, CB, VK, MJ, ENS, SA, and ACT supported data collection. JMP and RD conducted data analysis. All authors contributed to critical revisions and approved the final manuscript.

## Acknowledgements

We thank the HopeNet cohort study participants, without whom this research would not be possible. We also thank members of the HopeNet study team for research assistance; in addition to the named study authors, HopeNet team members who contributed to data collection and/or study administration during all or any part of the study were as follows: Phionah Ahereza, Dickson Beinomugisha, Patrick Gumisiriza, Justus Kananura, Allen Kiconco, Michael Matte, Patrick Lukwago Muleke, and Elizabeth Namara. We also thank Roger Hofmann of West Portal Software Corporation (San Francisco, CA, USA), for developing and customizing the Computer Assisted Survey Information Collection Builder software program used to collect the survey data.

## Data Available

Data and code for analysis are available from the corresponding author upon reasonable request.

## Ethics approval

Ethical approval was granted by the Mass General Brigham Institutional Review Board, the Research Ethics Committee at Mbarara University of Science and Technology, and the Vanderbilt Human Research Protections Program. We also received clearance from the Uganda National Council of Science and Technology and the Research Secretariat in the Office of the President of the Republic of Uganda. The analyses were not pre-registered; therefore, the results should be considered exploratory.

---

**Supplemental Table 1.** Perceived norms about male circumcision in own village among adult **<u>men</u>** across eight villages in Rwampara District, southwest Uganda (N=698).

|  | N of male<br>study<br>participants | n and percent of<br>men who believed<br>most men are<br>circumcised |  | n and percent of<br>men who<br>believed some<br>men are<br>circumcised |  | n and percent of<br>men who believed<br>few men are<br>circumcised |  | n and percent of<br>men did not<br>know how many<br>men are<br>circumcised |  |
| --- | --- | --- | --- | --- | --- | --- | --- | --- | --- |
| Total | 698 | 111 | 16% | 330 | 47% | 189 | 27% | 67 | 10% |
| Age (years) |  |  |  |  |  |  |  |  |  |
| 17-25 | 102 | 29 | 28% | 46 | 45% | 20 | 20% | 7 | 7% |
| 26-35 | 164 | 25 | 15% | 92 | 56% | 39 | 24% | 8 | 5% |
| 36-45 | 157 | 21 | 13% | 71 | 45% | 55 | 35% | 10 | 6% |
| 46-55 | 142 | 17 | 12% | 72 | 51% | 37 | 26% | 15 | 11% |
| ≥56 | 130 | 17 | 13% | 49 | 38% | 38 | 29% | 26 | 20% |
| Marital status |  |  |  |  |  |  |  |  |  |
| Not married and not<br>cohabiting | 205 | 40 | 20% | 93 | 45% | 50 | 24% | 22 | 11% |
| Married / cohabiting as if<br>married | 493 | 71 | 14% | 237 | 48% | 139 | 28% | 45 | 9% |
| Religion |  |  |  |  |  |  |  |  |  |
| Catholic | 155 | 22 | 14% | 75 | 48% | 40 | 26% | 17 | 11% |
| Muslim | 9 | 2 | 22% | 5 | 56% | 2 | 22% | 0 | 0% |
| Protestant | 505 | 83 | 16% | 234 | 46% | 140 | 28% | 48 | 10% |
| Other (Not religious; Seventh-Day Adventist; Born-again<br>Pentecostal) | 29 | 4 | 14% | 16 | 55% | 7 | 24% | 2 | 7% |
| Education |  |  |  |  |  |  |  |  |  |
| None / some primary<br>education | 224 | 31 | 14% | 98 | 44% | 65 | 29% | 29 | 13% |
| Completed primary education or more | 474 | 80 | 17% | 232 | 49% | 124 | 26% | 38 | 8% |
| Household asset wealth |  |  |  |  |  |  |  |  |  |
| 1st quintile (poorest) | 111 | 17 | 15% | 55 | 50% | 33 | 30% | 6 | 5% |
| 2nd quintile | 138 | 23 | 17% | 64 | 46% | 41 | 30% | 10 | 7% |
| 3rd quintile | 142 | 23 | 16% | 73 | 51% | 31 | 22% | 15 | 11% |
| 4th quintile | 152 | 24 | 16% | 68 | 45% | 39 | 26% | 20 | 13% |
| 5th quintile (least poor) | 155 | 24 | 15% | 70 | 45% | 45 | 29% | 16 | 10% |
| Lived with at least one woman who thought >50% of men in village had been circumcised |  |  |  |  |  |  |  |  |  |
| No | 559 | 91 | 16% | 263 | 47% | 151 | 27% | 54 | 10% |
| Yes | 139 | 20 | 14% | 67 | 48% | 38 | 27% | 13 | 9% |
| Had been tested for HIV in past 12 months |  |  |  |  |  |  |  |  |  |
| No | 383 | 57 | 15% | 175 | 46% | 106 | 28% | 44 | 12% |
| Yes | 315 | 54 | 17% | 155 | 49% | 83 | 26% | 23 | 7% |
| Had an STI in past 12 months |  |  |  |  |  |  |  |  |  |
| No | 637 | 96 | 15% | 299 | 47% | 176 | 28% | 65 | 10% |
| Yes | 40 | 9 | 23% | 23 | 58% | 8 | 20% | 0 | 0% |
| Had condomless sex with a non-spouse partner in past 12 months |  |  |  |  |  |  |  |  |  |
| No | 580 | 88 | 15% | 269 | 46% | 166 | 29% | 56 | 10% |
| Yes | 97 | 17 | 18% | 53 | 55% | 18 | 19% | 9 | 9% |
| Perceived personal HIV risk |  |  |  |  |  |  |  |  |  |
| HIV-positive | 67 | 12 | 18% | 27 | 40% | 22 | 33% | 6 | 9% |
| No/low risk | 546 | 90 | 16% | 267 | 49% | 138 | 25% | 51 | 9% |
| Medium/high risk | 77 | 9 | 12% | 33 | 43% | 26 | 34% | 8 | 11% |

**Supplemental Table 2.** Perceived norms about male circumcision in own village among adult **<u>women</u>** across eight villages in Rwampara District, southwest Uganda (N=868).

|  | N of female<br>study<br>participants | n and percent of<br>women<br>who believed<br>most men are<br>circumcised |  | n and percent of<br>women who<br>believed some<br>men are<br>circumcised |  | n and percent of<br>women<br>who believed few<br>men are<br>circumcised |  | n and percent of<br>women did not<br>know how many<br>men are<br>circumcised |  |
| --- | --- | --- | --- | --- | --- | --- | --- | --- | --- |
| Total | 868 | 164 | 19% | 301 | 35% | 177 | 20% | 220 | 26% |
| Age (years) |  |  |  |  |  |  |  |  |  |
| 18-25 | 139 | 32 | 23% | 40 | 29% | 37 | 27% | 30 | 22% |
| 26-35 | 213 | 40 | 19% | 89 | 42% | 48 | 23% | 36 | 17% |
| 36-45 | 182 | 31 | 17% | 72 | 40% | 41 | 23% | 38 | 21% |
| 46-55 | 146 | 38 | 26% | 55 | 38% | 23 | 16% | 29 | 20% |
| ≥56 | 174 | 21 | 12% | 42 | 24% | 28 | 16% | 78 | 46% |
| Marital status |  |  |  |  |  |  |  |  |  |
| Not married and not cohabiting | 337 | 56 | 17% | 97 | 29% | 71 | 21% | 109 | 33% |
| Married / cohabiting as if<br>married | 531 | 108 | 20% | 204 | 38% | 106 | 20% | 111 | 21% |
| Religion |  |  |  |  |  |  |  |  |  |
| Catholic | 189 | 31 | 16% | 75 | 40% | 39 | 21% | 44 | 23% |
| Muslim | 13 | 5 | 38% | 2 | 15% | 2 | 15% | 4 | 31% |
| Protestant | 617 | 120 | 19% | 211 | 34% | 123 | 20% | 157 | 26% |
| Other (Not religious; Seventh-Day Adventist; Born-again<br>Pentecostal) | 49 | 8 | 16% | 13 | 27% | 13 | 27% | 15 | 31% |
| Education |  |  |  |  |  |  |  |  |  |
| None / some primary education | 402 | 74 | 18% | 134 | 33% | 63 | 16% | 126 | 32% |
| Completed primary education or<br>more | 466 | 90 | 19% | 167 | 36% | 114 | 24% | 94 | 20% |
| Household asset wealth |  |  |  |  |  |  |  |  |  |
| 1st quintile (poorest) | 202 | 39 | 19% | 60 | 30% | 40 | 20% | 62 | 31% |
| 2nd quintile | 175 | 33 | 19% | 68 | 39% | 36 | 21% | 35 | 20% |
| 3rd quintile | 172 | 35 | 20% | 62 | 36% | 32 | 19% | 43 | 25% |
| 4th quintile | 161 | 29 | 18% | 54 | 34% | 37 | 23% | 40 | 25% |
| 5th quintile (least poor) | 158 | 28 | 18% | 57 | 36% | 32 | 20% | 40 | 25% |
| Lived with at least one woman<br>who thought >50% of men in<br>village had been circumcised |  |  |  |  |  |  |  |  |  |
| No | 643 | 22 | 3% | 271 | 42% | 154 | 24% | 190 | 30% |
| Yes | 225 | 142 | 63% | 30 | 13% | 23 | 10% | 30 | 13% |
| Had been tested for HIV in past 12<br>months |  |  |  |  |  |  |  |  |  |
| No | 416 | 83 | 20% | 134 | 32% | 65 | 16% | 130 | 32% |
| Yes | 452 | 81 | 18% | 167 | 37% | 112 | 25% | 90 | 20% |
| Had an STI in past 12 months |  |  |  |  |  |  |  |  |  |
| No | 803 | 153 | 19% | 285 | 35% | 157 | 20% | 202 | 25% |
| Yes | 35 | 6 | 17% | 9 | 26% | 8 | 23% | 12 | 34% |
| Had condomless sex with a non-spouse partner in past 12 months |  |  |  |  |  |  |  |  |  |
| No | 756 | 143 | 19% | 272 | 36% | 148 | 20% | 187 | 25% |
| Yes | 82 | 16 | 20% | 22 | 27% | 17 | 21% | 27 | 33% |
| Perceived personal HIV risk |  |  |  |  |  |  |  |  |  |
| HIV-positive | 113 | 25 | 22% | 43 | 38% | 24 | 21% | 21 | 19% |
| No/low risk | 598 | 101 | 17% | 202 | 34% | 126 | 21% | 164 | 28% |
| Medium/high risk | 143 | 35 | 24% | 48 | 34% | 26 | 18% | 33 | 23% |

**Supplemental Table 3.** Modified multivariable Poisson regression model estimating associations between the perceived norm about circumcision status among men in one’s village and being personally circumcised among almost all resident adult men across eight villages in Rwampara District, southwestern Uganda (n=649 men).

|  | Yes, circumcised |  |  |
| --- | --- | --- | --- |
|  | aRR | (95% CI) | p-value |
| Perceived norm about male circumcision uptake in own village |  |  |  |
| Most men are circumcised (i.e., >50%) | 1.82 | (1.45 – 2.28) | <0.001 |
| Some men are circumcised (i.e., 10% to <50%) | REF | - | - |
| Few men are circumcised (i.e., 0 to <10%) | 0.52 | (0.35 – 0.77) | 0.001 |
| Don't know how many men are circumcised | 0.73 | (0.44 – 1.21) | 0.221 |
| Age (years) |  |  |  |
| 18-25 | REF | - | - |
| 26-35 | 0.96 | (0.75 – 1.23) | 0.726 |
| 36-45 | 0.84 | (0.67 – 1.05) | 0.127 |
| 46-55 | 0.52 | (0.39 – 0.69) | <0.001 |
| 56+ | 0.23 | (0.15 – 0.35) | <0.001 |
| Married / cohabiting (vs. other) | 1.08 | (0.88 – 1.34) | 0.448 |
| Religion |  |  |  |
| Catholic | REF | - | - |
| Muslim | 2.85 | (2.13 – 3.81) | <0.001 |
| Protestant | 0.87 | (0.60 – 1.25) | 0.454 |
| Other | 1.31 | (0.83 – 2.07) | 0.241 |
| Completed primary education or more (vs. did not) | 1.64 | (1.13 – 2.39) | 0.009 |
| Household asset quintile |  |  |  |
| 1st quintile (poorest) | REF | - | - |
| 2nd quintile | 0.95 | (0.69 – 1.29) | 0.734 |
| 3rd quintile | 0.88 | (0.53 – 1.45) | 0.604 |
| 4th quintile | 0.94 | (0.67 – 1.30) | 0.688 |
| 5th quintile (least poor) | 1.24 | (0.85 – 1.82) | 0.269 |
| Lived with at least one woman who thought >50% of men in village had been circumcised (vs. did not) | 1.10 | (0.71 – 1.72) | 0.669 |
| Tested for HIV in past 12 months (vs. did not) | 1.09 | (0.82 – 1.46) | 0.539 |
| Had an STI in past 12 months (vs. did not) | 1.01 | (0.54 – 1.92) | 0.964 |
| Had condomless sex with a non-spouse partner in past 12 months (vs. did not) | 1.07 | (0.88 – 1.31) | 0.492 |
| Perceived personal HIV risk |  |  |  |
| HIV-positive | 1.13 | (0.74 – 1.74) | 0.565 |
| No/low risk | REF | - | - |
| Medium/high risk | 1.03 | (0.90 – 1.17) | 0.710 |

aRR = adjusted relative risk ratio, CI = confidence interval, REF = reference group

